# Circulating PACAP levels are associated with altered imaging measures of entorhinal cortex neurite density in posttraumatic stress disorder

**DOI:** 10.1101/2023.08.31.23294894

**Authors:** Steven J Granger, Victor May, Sayamwong E Hammack, Eylül Akman, Sydney A. Jobson, Elizabeth A Olson, Cameron D Pernia, Nikos P Daskalakis, Caitlin Ravichandran, William A Carlezon, Kerry J Ressler, Scott L Rauch, Isabelle M Rosso

**Affiliations:** Division of Depression and Anxiety Disorders, McLean Hospital, Belmont, MA; Basic Neuroscience Division, McLean Hospital, Belmont, MA; Department of Pediatrics, Harvard Medical School, Boston, MA; Department of Psychiatry, Harvard Medical School, Boston, MA; Larner College of Medicine, University of Vermont, Burlington, VT; Lurie Center for Autism, Massachusetts General Hospital, Lexington, MA; Stanley Center for Psychiatric Research, Broad Institute of MIT and Harvard, Cambridge, MA

**Keywords:** posttraumatic stress disorder, diffusion imaging, medial temporal lobe, pituitary adenylate cyclase-activating polypeptide, stress peptide, entorhinal cortex

## Abstract

**Background:** Pituitary adenylate cyclase-activating polypeptide (PACAP) regulates plasticity in brain systems underlying arousal and memory and is associated with posttraumatic stress disorder (PTSD). Research in animal models suggests that PACAP modulates entorhinal cortex (EC) input to the hippocampus, contributing to impaired contextual fear conditioning. In PTSD, PACAP is associated with higher activity of the amygdala to threat stimuli and lower functional connectivity of the amygdala and hippocampus. However, PACAP-affiliated structural alterations of these regions have not been reported. Here, we examined whether peripheral PACAP levels were associated with neuronal morphology of the amygdala and hippocampus (primary analysis), and EC (secondary analysis) using Neurite Orientation Dispersion and Density Imaging.

**Methods:** Sixty-four (44 female) adults (19 to 54 years old) with DSM-5 Criterion A trauma exposure completed the Clinician-Administered PTSD Scale (CAPS-5), a blood draw, and magnetic resonance imaging. PACAP38 radioimmunoassay was performed and T1-weighted and multi-shell diffusion-weighted images were acquired. Neurite Density Index (NDI) and Orientation Dispersion Index (ODI) were quantified in the amygdala, hippocampus, and EC. CAPS-5 total score and anxious arousal score were used to test for clinical associations with brain structure.

**Results:** Higher PACAP levels in blood were associated with greater EC NDI (β=0.31, q=0.034) and lower EC ODI (β=-0.30, q=0.042) and not hippocampal or amygdala measures. Neither EC NDI nor ODI was associated with clinical measures.

**Conclusions:** Circulating PACAP levels were associated with altered neuronal density of the EC but not hippocampus or amygdala. These findings strengthen evidence that PACAP may impact arousal-associated memory circuits.

## Introduction

Posttraumatic stress disorder (PTSD) is characterized by intrusion, avoidance, negative emotions, and hyperarousal symptoms after exposure to severe trauma (1). Pituitary adenylate cyclase-activating polypeptide (PACAP) is a well-established neuromodulator of stress and arousal (2), is involved in the secretion and production of corticotrophin-releasing hormone (3–5), and has been associated with hyperarousal symptoms of PTSD (6). Stress-induced morphological alterations of the medial temporal lobe (MTL), including changes in amygdala and hippocampal volume, are implicated in PTSD (7).

Notably, circulating PACAP levels and PAC1 receptors (PAC1Rs, encoded by *ADCYAP1R1*) have been associated with modulation of MTL functioning in animal models (8), healthy adults (9), and people with PTSD (10). Research in animal models has shown that PACAP-regulated pathways influence neuronal differentiation and are involved in neurotrophic processes including synaptogenesis and axon outgrowth in stress-sensitive MTL regions (11,12). Others have suggested that an *ADCYAP1R1-*associated risk polymorphism increases hippocampal vulnerability to the harmful effects of stress (10). However, a relationship between PACAP and MTL morphometry has not been reported in PTSD. To bridge this research gap, the recent emergence of neurite orientation dispersion and density imaging (NODDI) presents a timely opportunity. NODDI provides metrics of the orientations of axons and dendrites (together known as “neurites”), which represent potentially functionally-relevant tissue features not discerned by traditional volumetric analyses. Consequently, this study investigated the association between PACAP and NODDI indices within MTL regions in a sample of adults with PTSD.

Research in animal models suggests that PACAP modulates multiple nodes of MTL circuitry, with implications for fear learning. Within the MTL, PAC1Rs are densely expressed in the amygdala, entorhinal cortex (EC), and hippocampus (8,13–15). Brief application of PACAP in rat hippocampal slices enhances synaptic activity and strength within the hippocampus and modulates fear conditioning (8,16,17). Similarly, PAC1R knockout mice exhibit deficits in long-term potentiation following high-frequency stimulation of EC input to the hippocampus (18,19). Within the amygdala, PACAP infusion prior to fear conditioning increases synaptic plasticity and neuronal activation in a manner that disrupts fear acquisition in the short-term and yet facilitates longer-term retention of fear cues (20). Altogether, this literature highlights the role of PACAP in modulating synaptic signaling, dendritic and axonal growth, and fear learning.

PACAP systems have also been implicated in arousal-related symptoms and underlying MTL functioning in PTSD. In a seminal report, Ressler and colleagues (2011) found that circulating PACAP, *ADCYAP1R1* SNP polymorphisms, and *ADCYAP1R1* DNA methylation were associated with PTSD diagnosis, and this has since been replicated in independent samples (21–23) but see (24) Circulating PACAP was also correlated with significantly greater severity of hyperarousal symptoms in women with PTSD (6). In a functional imaging study of traumatized women, *ADCYAP1R1* polymorphism predicted greater amygdala reactivity to fearful stimuli and lower functional connectivity between the amygdala and hippocampus (10). Similarly, our group has recently shown that greater circulating PACAP was related to greater amygdala connectivity with posterior default mode network regions (25). Together, this motivates investigation of PACAP-affiliated MTL structural indices that may influence MTL functional abnormalities.

Multiple lines of research support investigating an association of PACAP with EC phenotypes in PTSD. The EC functions as a gateway for amygdala modulation of hippocampal activity (26). As an intermediate node, the EC may be an additional factor impacting the previously reported associations of PACAP with amygdala and hippocampus functional connectivity. This suggestion is particularly relevant considering PACAP-affiliated glutamatergic neurons of the EC that project to the hippocampus (8). The EC also supports threat processing functions that are known to be modulated by PACAP (27,28), including contextual fear conditioning (27,29). However, prior studies did not examine whether PACAP relates to structural characteristics of the EC in humans.

Diffusion-weighted MRI, specifically NODDI (30), has been used to detect differences in gray matter microstructure. NODDI provides researchers with surrogate measures of neuronal integrity hypothesized to relate to the number of neurites and complexity of dendrites (Neurite Density Index: NDI) as well as the dispersion of axons and neurons (Orientation Dispersion Index: ODI) which have been shown to be related to functional dynamics (31). NODDI metrics have recently been proposed as candidates for next-generation biomarkers in psychiatry (32,33); however, to our knowledge these methods have never been applied to study structural brain correlates of the stress response in individuals with PTSD. Using NODDI could enhance understanding of cellular characteristics, differences, or changes in PTSD that might influence function.

To address this gap in the literature, this study examined whether peripheral PACAP was associated with regional NODDI measures of gray matter microstructure in the MTL of adults with DSM- 5 Criterion A trauma exposure and a range of PTSD symptoms. Our primary analyses examined PACAP in relation to amygdala and hippocampus NDI and ODI, given previous findings of altered activity of amygdala and connectivity with the hippocampus in individuals with the *ADCYAP1R1* polymorphism genotype. As a secondary and exploratory analysis, we examined whether peripheral PACAP was associated with NDI and ODI of the EC, based on evidence that it serves as an intermediary region between the hippocampus and amygdala. To determine the clinical relevance of significant findings, we examined whether PACAP-associated NDI and ODI metrics were related to total PTSD symptom severity and anxious arousal symptoms. Examination of anxious arousal symptoms was based on previous evidence that higher peripheral PACAP levels were associated with hyperarousal symptoms and that individuals with a polymorphism in the gene encoding PAC1Rs may show greater physiological fear responses including hyperarousal and startle discrimination (6). Factor analytic investigations of the latent structure of PTSD symptoms have shown that self-reported hypervigilance and startle symptoms cluster together as a part of an anxious arousal construct, separately from symptoms of dysphoric arousal (e.g., irritability, trouble sleeping) (34).

## Methods and Materials

### Participants

One-hundred and three trauma-exposed community-based adults were recruited from the greater Boston metropolitan area. Of these individuals, eighty-four were scanned with the Human Connectome Project (HCP) Adult Lifespan protocol and nineteen with the HCP Young Adult protocol. There is evidence that NODDI metrics across gray and white matter are susceptible to differences in acquisition methods. For instance, NDI is sensitive to the choice of outer shell b-value, while ODI is sensitive to the number of gradient directions (35), and others have shown greater variability of NODDI measures in shell-schemes with low b-values (36). To avoid the possibility that NODDI measures are impacted by varying protocols in this clinically-assessed sample, we used only the participants scanned with the majority-use HCP Lifespan protocol. Twenty of the eighty-four individuals scanned with the HCP Adult Lifespan protocol did not have processed PACAP data. The final sample consisted of sixty-four trauma-exposed community-based adults (19 to 54 years old; 44 female). Participants were included if they met DSM-5 Criterion A trauma exposure for PTSD and at least two of the four PTSD symptom clusters (B-E). Exclusion criteria were left-handedness, confounding medical condition including untreated seizure disorder or neurological disorder, inability to tolerate blood draws, history of head trauma with loss of consciousness > 5 min, current treatment with an antipsychotic (unless prescribed for PTSD-related sleep and nightmares), current (past month) alcohol use disorder or moderate/severe substance use disorder, current psychotic disorder, anorexia, obsessive-compulsive disorder, manic or mixed mood episode, lifetime history of schizophrenia or schizoaffective disorder, MR contraindications (e.g., metal implants and claustrophobia), positive pregnancy test on day of MRI scan (female participants), and history of receiving hormonal replacement therapy or undergoing gender confirmation surgery. All study procedures were approved by the Mass General Brigham Human Research Committee (Protocol 2019P000626). Characteristics of the sample are summarized in Table 1.

**Table 1.**
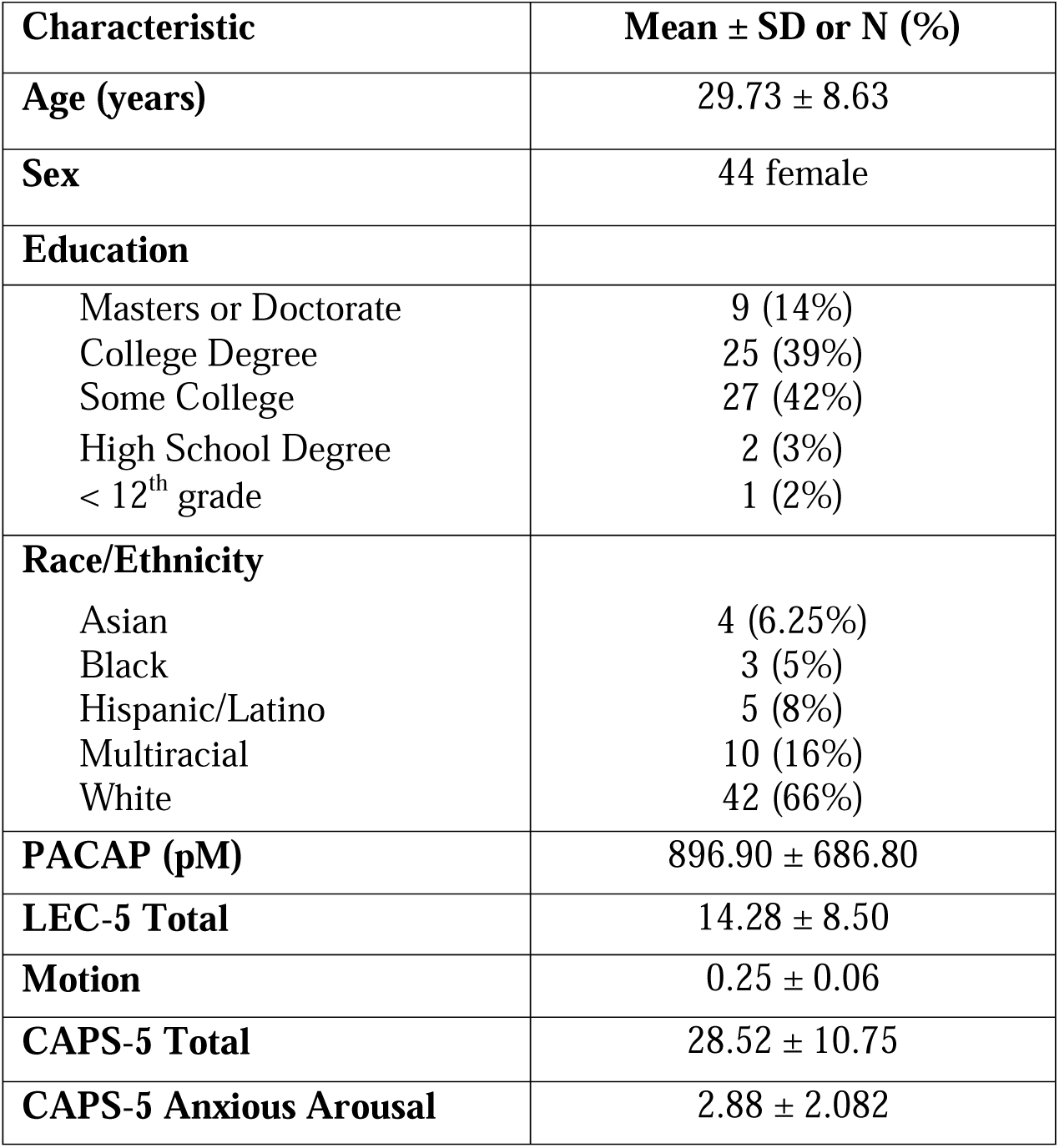
Sample Characteristics (N = 64) Note: PACAP = pituitary adenylate cyclase-activating polypeptide; LEC-5 Total = Life Events Checklist for DSM-5 total; motion = average relative motion during scan; CAPS-5 = Clinician Administered PTSD Scale for DSM-5

### Clinical Interviews and Questionnaires

#### Life Events Checklist (LEC-5)

The LEC-5 was given as a survey of 17 potentially traumatic events and used to determine DSM- 5 Criterion A trauma exposure and an index trauma for the interview.

#### Clinician-Administered PTSD Scale for DSM-5 (CAPS-5)

The CAPS-5 (37) was administered by doctoral-level psychologists and used to determine DSM- 5 PTSD diagnosis and symptom severity. We derived CAPS-5 total score and anxious arousal subscore. The latter equaled the sum of responses to Item 17 (E3) Hypervigilance and Item 18 (E4) Exaggerated Startle Response (34).

### PACAP Immunoassays

Blood samples were collected at the beginning of each study visit, between approximately 8:00 AM -12:30 PM. Participants were instructed not to eat the morning of their visit prior to their blood draw. Human plasma samples were prepared as described previously (6,38), and all human PACAP38-specific measurements were performed at the University of Vermont, Larner College of Medicine, using double antibody sandwich ELISA immunoassays (Cat. No. HUFI02692, AssayGenie, Dublin, Ireland). Samples were centrifuged at 3500 rpm for 15 minutes. Plasma was extracted and stored at -80 °C until analysis.

Optimal sample volume was determined in dilution tests, and all values represent the mean from assay duplicates; intra-assay variation was approximately 9%. The assay midpoint was 1.1 fmol and the detection limit from the linear range of the standard curve was 0.2 fmol. Outliers with exceedingly high (n = 2) concentrations of PACAP levels were winsorized to the next highest reliable/non-outlier value (38).

### Imaging

MRI data were acquired on a 3.0 Tesla Siemens Prisma Scanner at the McLean Imaging Center. Anatomical whole-brain images were obtained. A T1-weighted MP-RAGE sequence was acquired; TR = 2500ms, TE = 7.27ms, flip angle = 8 deg, FOV = 256 x 256, voxel size = 0.8mm isotropic. A high-angular multi-shell whole brain diffusion-weighted imaging sequence was also acquired; TR = 3230, TE = 89.5ms; flip angle = 78deg, refocusing flip angle = 160deg, FOV = 210 x 210; matrix = 168 x 144, slice thickness = 1.5mm, 92 slices, voxel size = 1.5mm isotropic, b = 1500 and 3000 s/mm^2^, ∼47 directions/shell, multiband factor = 4. An identical sequence acquired with opposite-phase encoding was also acquired and used to correct for magnetization-induced susceptibility distortion (see below).

### Image Processing

T1-weighted images were processed using Freesurfer6 (39). Freesurfer-processed T1-weighted images were skull-stripped using AFNIs 3dSkullStrip program and corrected for signal intensity changes (40). Opposite phase encoding B0’s were merged for FSL’s topup program (41,42). Topup results were fed into FSL’s eddy program to correct for motion, eddy current artifacts, and magnetization induced susceptibility distortion (43). We used the –repol option to replace outliers and further improve the quality of the data (44). Rotated vectors were used for subsequent analysis. Neurite Density Index (NDI) and Orientation Dispersion Index (ODI) were calculated using the Microstucture Diffusion Toolbox (MDT) (45). Skull-stripped T1 data were aligned to the NDI image using antsRegistrationSynQuick.sh from Advanced Normalization Tools (ANTS) (46). Diffeomorphic transformations from the skull-stripped T1 image to the NDI image were applied to each region of interest using ANTS. Voxel data for each hemisphere, measure, and region type were extracted using AFNI’s 3dmaskdump (40). Alignment of each region of interest to the NDI image was checked with tools distributed with scikit-learn (47). We analyzed histograms of raw voxel values for all metrics bilaterally. We observed a non-normal distribution of voxel intensities in NDI metrics. To correct this bias, we filtered out voxels with NDI values of over 0.98. All individuals had at least 70% retained voxels after voxel filtering (Supplementary Table 1). Participant motion was quantified using FSLs quality control eddy_quad program for average motion relative to the previous sub-brick and was included as a nuisance regressor in regression models (48).

### Statistical Analyses

Linear regressions were used to examine circulating PACAP levels in relation to NODDI metrics of primary interest (NDI and ODI from the amygdala and hippocampus) and secondary interest (NDI and ODI from the EC), controlling for sex, age, and participant motion. Estimates of both standardized and unstandardized regression coefficients were calculated to aid the interpretation of effect sizes. For regional NODDI metrics that were significantly associated with PACAP after multiple comparison correction, post hoc linear regression examined their associations with PTSD symptom severity, as quantified by CAPS-5 total score and anxious arousal subscore, controlling for age, sex, and motion.

Hommel multiple testing corrections were applied separately to hypothesis testing results for models quantifying associations of primary interest (4 models; amygdala and hippocampal NDI and ODI) and quantifying associations of secondary interest (2 models; EC NDI and ODI). No multiple testing correction was applied to results of post hoc analyses. Data analysis was conducted in R-Studio (http://r-project.org/.). Confidence intervals were calculated at the individual 95% confidence level using the confint function from R. Standardized beta scores were calculated using R-Studio’s lm.beta package (49). Statistical significance tests were two-sided and conducted at the alpha=0.05 significance level after correction.

## Results

Table 2 shows the results of all multiple linear regression analyses and the unstandardized and standardized coefficient estimates for all covariates.

**Table 2.** Results of multiple linear regression for our primary analyses (models predicting hippocampal and amygdala NDI and ODI in bold) and secondary analyses (models predicting entorhinal cortex NDI and ODI) Note: Multiple Linear Regression results for PACAP, age, sex, and motion predicting NDI and ODI for each region. q-values (corrected p-values) are reported. We corrected for multiple comparisons using Hommel for our primary (amygdala and hippocampus; shown in bold) and secondary analyses (entorhinal cortex) separately. Unadjusted beta values were multiplied by 1000 for ease of reporting. Unadjusted 95% confidence intervals are reported. * Indicates significance at the corrected level, ┼ indicates significance at the uncorrected level.

|  | NDI x 1000 |  |  | ODI x 1000 |  |  |
| --- | --- | --- | --- | --- | --- | --- |
| | B (95% C.I.) | $\beta$ | q | B (95% C.I.) | $\beta$ | q |
| *PACAP* |  |  |  |  |  |  |
| **Amygdala** | **0.00411 (-0.00235, 0.0106)** | **0.17** | **0.61** | **0.00253 (-0.00723, 0.0123)** | **0.075** | **0.61** |
| **Hippocampus** | **0.00680 (0.000641, 0.0130)** | **0.27** | **0.12<sup>+</sup>** | **0.00257 (-0.00534, 0.0105)** | **0.095** | **0.61** |
| Entorhinal Cortex | 0.00999 (0.00187, 0.0181) | 0.31 | 0.034* | -0.00748 (-0.0147, -0.000280) | -0.30 | 0.042* |
| *Age* |  |  |  |  |  |  |
| Amygdala | 0.81 (0.33, 1.30) | 0.41 | 0.005* | 0.363 (-0.374, 1.099) | 0.14 | 0.59 |
| Hippocampus | 0.73 (0.27, 1.20) | 0.37 | 0.008* | 0.163 (-0.434, 0.760) | 0.076 | 0.59 |
| Entorhinal Cortex | 0.855 (0.24, 1.47) | 0.34 | 0.014* | 0.383 (-0.160, 0.926) | 0.19 | 0.16 |
| *Sex* |  |  |  |  |  |  |
| Amygdala | 5.96 (-4.05, 16.0) | 0.16 | 0.51 | -1.21 (-1.62, 1.39) | -0.025 | 0.87 |
| Hippocampus | 12.9 (3.39, 22.5) | 0.35 | 0.035* | 5.88 (-6.37, 18.1) | 0.15 | 0.68 |
| Entorhinal Cortex | 10.3 (-2.27, 22.88) | 0.22 | 0.22 | -5.43 (-16.571, 5.714) | -0.15 | 0.33 |
| *Motion* |  |  |  |  |  |  |
| Amygdala | -6.21 (-68.7, 56.3) | -0.023 | 0.86 | 55.1 (-39.3, 149) | 0.15 | 0.86 |
| Hippocampus | -5.30 (-64.9, 54.3) | -0.019 | 0.86 | -29.2 (-105.7, 47.4) | -0.098 | 0.86 |
| Entorhinal Cortex | 31.70 (-46.92, 110.25) | 0.091 | 0.51 | -22.9 (-92.473, 46.752) | -0.083 | 0.51 |

### Primary Analysis: Relationship of circulating PACAP levels with Hippocampus and Amygdala NODDI Metrics

In the regression examining circulating PACAP levels in relation to hippocampal NDI, there was a positive effect of PACAP at the uncorrected level (β = 0.27, p = 0.031), which did not survive correction for multiple comparisons. Hippocampal NDI was positively correlated with age (β = 0.37, q = 0.008) and was greater in males (β = 0.35, q = 0.035). Hippocampal NDI was not related to participant motion. In the regression of PACAP on hippocampal ODI, the effect of PACAP was not significant, nor were the effects of the age, sex, and motion covariates.

The regression of PACAP and amygdala NDI was not significant, nor were the contributions of sex and participant motion. However, we observed a significant positive effect of age on amygdala NDI (β = 0.41, q = 0.005). In the regression of PACAP and amygdala ODI, the effects of PACAP, age, sex, and participant motion were all non-significant.

### Secondary Analysis: Relationship of PACAP with EC NODDI Metrics

In the regression of PACAP and EC NDI, there was a significant positive effect of PACAP at the uncorrected and corrected levels (β = 0.31, q = 0.034), controlling for a significant positive effect of age (β = 0.34, q = 0.014) and non-significant effects of sex and participant motion. In the regression of PACAP and EC ODI, there was a significant negative effect of PACAP at the uncorrected and corrected levels (β = -0.30, q = 0.042), controlling for non-significant effects of age, sex, and participant motion.

### Post hoc Correlational Analyses of NODDI Metrics with Clinical Symptoms

Neither EC NDI (β = 0.011, p = 0.94) nor ODI (β = -0.19, p = 0.13) was associated with CAPS-5 total score, covarying for age, sex, and motion. In addition, neither EC NDI (β = 0.13, p = 0.36) nor ODI (β = -0.11, p = 0.42) was associated with the anxious arousal subscore, covarying for age, sex, and motion.

## Discussion

Here we examined the relationship between PACAP levels circulating in blood and NODDI measures of dendritic and cellular complexity of the MTL in people with PTSD. Our findings suggest that circulating PACAP is particularly relevant to the microstructure of the EC. We found that higher PACAP levels were associated with higher NDI and lower ODI of the EC, after accounting for age, sex, and motion. While we had stronger a priori predictions regarding the associations between PACAP levels and the microstructure of the amygdala and hippocampus, a positive association between PACAP levels and hippocampal NDI was not significant after correcting for multiple comparisons. Finally, exploration of symptom correlations revealed that EC NODDI measures did not track with total PTSD symptom severity or anxious arousal symptoms. Altogether, these findings suggest that higher levels of peripheral PACAP are associated with EC microstructural properties in PTSD.

Our strongest finding indicates that higher PACAP levels are associated with greater NDI of the EC, which may reflect lower dendritic complexity (50). Studies of aging have shown that greater hippocampal NDI is found in older adults compared with younger adults, and is associated with poorer episodic memory performance, thus capturing morphological changes related to impaired MTL gray matter function (50,51). Our findings of positive associations between age and NDI of the amygdala, hippocampus, and EC are consistent with this literature on aging, lending confidence that they may reflect similar underlying microstructural properties. Notably, PACAP enhances the excitability of the hippocampus through upstream regions such as the EC (8) and via known projections (52,53). Using magnetic resonance spectroscopy, we previously found lower neuronal integrity along with higher glutamate levels in the hippocampus of individuals with PTSD, which is potentially compatible with excitotoxic mechanisms (54). Therefore, we speculate that a reduction in dendritic complexity of the EC in association with PACAP may have downstream consequences on stress-related excitotoxic-neuronal loss in the hippocampus. Further research is needed to fully test this hypothesis in relation to PACAP.

PACAP was also associated with lower EC ODI, which indicates less occupancy of the extraneurite space (33). Animal studies pairing immunohistochemistry and high-resolution diffusion-weighted MRI suggest that reduced ODI in gray matter is associated with selective depletion of microglia (55). Changes in glial cell occupancy may drive lower ODI values found in gray matter, as glia take up a large proportion of extra-neurite space. Hence, we speculate that higher peripheral PACAP may be associated with lower microglial density (lower ODI) in the EC in PTSD. Altogether, these associations between PACAP and altered NODDI measures of the MTL have implications for studies that have found differences in EC volume (56–58) and function (59) in individuals with PTSD.

The observation that circulating PACAP levels are associated with altered EC NODDI measures is important in the context of prior research showing altered connectivity between the amygdala and hippocampus in trauma-exposed individuals with the *ADCYAP1R1* genotype (10). Because the EC acts as a gateway for amygdala influence on hippocampus-based functions (26), we suggest that PACAP may induce morphological changes in the EC that hinder effective communication between the amygdala and hippocampus. This suggestion may be likely given that alterations in NODDI measures are related to resting state connectivity dynamics (31). However, future research is needed to test the hypothesis that structural changes of the EC may act as an intermediate outcome influencing the relationship between PACAP and lower functional connectivity of the amygdala and hippocampus.

Unexpectedly, this investigation did not reveal a relationship between PACAP and NDI and ODI of the hippocampus and amygdala, potentially suggesting that the structural impact of PACAP is specific to the EC. This is surprising given our prior work in animals showing that PACAP signaling in the central nucleus of the amygdala increases anxiety-like responses (60) and our work in humans showing higher amygdala reactivity to threat stimuli and lower amygdala-hippocampus functional connectivity in women with PACAP-receptor genetic polymorphism (10). One potential reason for our inability to detect differences within the amygdala could be the specificity of action of PACAP at the amygdala subnuclei, which we did not investigate in this study.

Along these lines, in the current investigation, we looked at the hippocampus as a unitary structure without regard to its nuanced subregional anatomy and function (61,62). We have previously shown evidence that high-resolution diffusion MRI can outperform whole-brain standard resolution in the detection of tensor-based microstructural differences between subregions of the MTL, including DG and CA1 of the hippocampus (63). Thus differences in microstructure of hippocampal subregions conducted with standard resolutions (i.e., 1.5mm isotropic) related to various pathologies may be less meaningful or could be improved with high-resolution diffusion imaging, as others have also recently shown (64). Due to this prior work, we were hesitant to pursue subregional analysis. An important future direction of this work would be to determine if higher circulating PACAP is associated with microstructural measurements of hippocampal subfields and if these relationships are improved with the use of high-resolution diffusion-weighted MRI. We also chose a unitary structure for the EC despite its domain-sensitive subregions specializing in different episodic memory processes (65–67). Parsing the medial and lateral EC with high-resolution diffusion imaging would be an important future direction, given their relevance to posterior-medial and anterior-temporal cortical networks, which may provide further insight into the neurobiological impact of greater circulating PACAP in arousal-mediated memory circuits (68,69). Despite the potential that increased granularity NODDI measures within these regions may yield improved information, to our knowledge, we are the first to apply this method to studies of MTL gray matter in PTSD.

Our results should be interpreted in the context of numerous strengths and limitations. Considering the lack of published work in which NODDI methods have been applied to the MTL in individuals with PTSD, interpreting the results is challenging. While it is possible that these imaging measures reflect less dendritic complexity and glial cell density, cell-type interpretations of NODDI measures must be made cautiously (70). Histological validation of the relationship between PACAP and EC NODDI metrics in animal models would provide additional biological inference of the underlying mechanisms. In addition, this study had a small sample size primarily composed of females. Because of prior work finding that the relationship between PACAP and hyperarousal was selective to females (6) and literature showing differential development of PACAP receptors in the female hippocampus (71), future work with larger sample sizes should seek to investigate the impact and interaction of sex, PACAP, and subfield structure. Future work should also investigate the link between circulating PACAP levels relative to brain PACAP levels. This investigation is also characterized by several strengths. We implemented opposite-phase encoding to correct for magnetization-induced susceptibility distortion, which is relevant given that the MTL is particularly susceptible to signal distortion (72). In addition, we included motion as a nuisance regressor and found that the relationship between PACAP and EC NDI and ODI persisted. Finally, we observed a non-normal distribution of NDI values across all voxels of the EC on a subject-by-subject basis. This necessitated filtering of problematic voxels that likely reflected CSF or pia/dura mater. As we initially noticed a very strong relationship between PACAP and EC and hippocampal NDI prior to filtering, the notion that these relationships persist upon removing voxels with abnormally high NDI strengthens our confidence in these findings.

In conclusion, our findings highlight the association between circulating PACAP levels and gray matter microstructure of the EC in individuals with PTSD. Higher PACAP levels were related to greater NDI and smaller ODI, suggesting less EC dendritic complexity and cellular density, respectively. These findings may have implications for understanding the cellular integrity of the MTL and motivate inquiry into the role that EC microstructure may play in PTSD-related alterations of the amygdala-hippocampus network. Future research examining this interpretation needs to include histological validation as a function of region and tissue type. In addition, future studies should examine subregions of the hippocampus and amygdala to further elucidate the role of PACAP within the MTL in PTSD.

## Data Availability

All data produced in the present study are available upon reasonable request to the authors

## Acknowledgments

This work was supported by NIH awards P50MH115874 (to WAC and KJR, PDs; IMR, SLR, Project 4 PIs), R01MH120400 (IMR), R01MH97988 (SEH and VM), and NM119467 (CR).

---

**Supplementary Table 1.** Summary of NDI voxel filtering. Summary of percent voxels remaining in region after removal of voxels with NDI > 0.98.

| | Average ± SD<br>Percent Retained<br>after filtering | Range Percent<br>Retained after<br>filtering |
| --- | --- | --- |
| Left Entorhinal<br>Cortex | 90.35 ± 6.93 | 70.40 - 99.65 |
| Right Entorhinal<br>Cortex | 92.52 ± 5.69 | 76.14 - 99.76 |
| Left Amygdala | 98.24 ± 2.22 | 90.49 – 100 |
| Right Amygdala | 98.52 ± 1.86 | 90.66 - 100.00 |
| Left Hippocampus | 95.69 ± 2.46 | 86.19 - 99.27 |
| Right<br>Hippocampus | 95.46 ± 1.96 | 88.16 - 98.62 |

